# Disproportionate High-Risk Nonsteroidal Anti-inflammatory Drug (NSAID) Prescribing in Rural Virginia

**DOI:** 10.1101/2025.01.03.25319965

**Authors:** Michelle S. Rockwell, Christopher Grubb, Jamie K. Turner, Matthew Vinson, Isaiah Yim, Alexandra L. Hanlon, John W. Epling

## Abstract

**Introduction:** Individuals living in rural areas experience limited access to safe and effective pain management. Using insurance claims for 1.3 million Virginians, we evaluated variation in high-risk nonsteroidal anti-inflammatory drug (NSAID) prescribing by rurality during 2019-2021.

**Methods:** We applied a difference-in-differences model to analyze the effect of the COVID-19 pandemic on high-risk NSAID prescribing rates, stratifying incidence rate ratios (IRR) by rurality.

**Results:** Although high-risk NSAID prescribing rates decreased modesty during 2020-2021, rural areas experienced significantly higher prescribing rates throughout the study period (IRR: 1.594 [95% CI: 1.408, 1.803], p<.001).

**Conclusions:** Context-driven efforts to de-implement high-risk NSAID prescribing in rural Virginia are needed.

## INTRODUCTION

More than 55 million Americans (25% of the adult population) experience chronic pain.(1) Nonsteroidal anti-inflammatory drugs (NSAIDs) are among the most common pain therapies but, unfortunately, pose potential harm to some individuals. The National Kidney Foundation, American Heart Association, American Geriatrics Society, and others recommend limiting or avoiding NSAIDs in most patients with chronic kidney disease (CKD), heart failure (HF), and hypertension (HTN) due to risk of adverse effects.(2–6) Despite these recommendations, undesirable and increasing rates of high-risk NSAID use persist.(7–9)

Understanding contextual factors that influence high-risk NSAID prescribing is crucial to informing the design and implementation of deprescribing interventions. Rurality is an underexplored contextual factor that may influence high-risk NSAID prescribing. Individuals living in rural areas are more likely to hold manual labor jobs, experience limited access to pain management specialists and nonpharmacological pain therapies, and encounter financial barriers to care, all factors that influence NSAID use.(10,11) Health system disruptions, such as those induced by the COVID-19 pandemic (declared by the World Health Organization in March 2020), may also impact medication prescribing patterns.

In this study, we evaluated the influence of two contextual factors – residential rurality and the COVID-19 pandemic – on prescription high-risk NSAID prescribing in Virginia during 2019-2021. We hypothesized that 1) high-risk NSAID prescribing rates were greater in rural vs. non-rural areas and 2) high-risk NSAID prescribing rates increased during the pandemic.

## METHODS

This retrospective cohort study used data from the Virginia All-Payer Claims Database (APCD), which includes insurance claims for more than 6 million Virginians covered by multiple public and private insurers. We limited this study to adult patients who had both medical and pharmaceutical claims filed for 2019, 2020, and 202. Based on the use of de-identified data, this study was deemed non-human subjects research by the Institutional Review Board of Carilion Clinic.

### Procedures

From aggregated, population-level APCD claims, we established a study cohort of patients who had continuous enrollment with commercial, Medicare Advantage, or Medicaid payers for ≥ 12 retrospective months during 2019, 2020, and 2021. Traditional Medicare and dual eligible patients were excluded due to unavailability of Medicare Part D claims.

Using the Milliman MedInsight Health Waste Calculator (v. 7.2), we identified claims for NSAID prescriptions filled by patients with a diagnosis of CKD, HF, and/or HTN on record during the previous 12 months, as described previously.(12,13) We categorized claims as *clinically-indicated* or *high-risk*. Claims for aspirin were considered clinically-indicated (to avoid misclassification of prophylactic use) as were those for topical NSAIDs. All other NSAID prescribed for patients with CKD, HF, and/or HTN were categorized as high-risk.

We calculated the bi-monthly prescribing rate per 1000 patients for high-risk and clinically-indicated NSAID prescription claims during 2019-2021. Results were stratified by rurality using Rural-Urban Commuting Area (RUCA) codes (RUCA 1-3= non-rural, 4-10= rural),(14) age, and biological sex. Gender data is not available in the APCD. We extrapolated high-risk NSAID prescribing results to the population of insured adults in Virginia (5.9 million).

### Statistical Analysis

To evaluate the effect of the pandemic on NSAID prescribing rates, we used a heterogeneous difference-in-differences (D-in-D) design, allowing the pandemic D-in-D effect to vary within subpopulations defined by rurality, age, and biological sex. Poisson regression models were fit for high-risk and clinically-indicated prescribing rates. Predicted prescribing rates were estimated using model predictions with the absence of the D-in-D effect. An autoregressive correlation structure (AR1) was implemented to account for the correlation between neighboring month-pairs and a linear year term and categorical month-pair terms were applied to account for annual and secular trends in both models. Resulting models were expressed using incidence rate ratios (IRRs) and interaction effects were calculated using predicted marginal means with population ratios matching the study sample. All statistical analyses were performed by CG using the geepack (v1.3.11) and emmeans (v1.10.2) libraries within R (v4.4.1).

## RESULTS

The study cohort included 1.3 million patients (mean age 57.6 years, 56% female, 15% rural) (**TABLE 1**). Demographic characteristics remained stable from 2019-2021, with the exception of insurer. Mirroring state and national trends, the proportion of patients covered by commercial insurers decreased, while Medicaid increased during 2019-2021.

**Table 1.** Demographic Characteristics of Study Cohort.

|  | **2019**<br>N=1,264,734<br>n(%) | **2020**<br>N=1,418,555<br>n(%) | **2021**<br>N=1,509,583<br>n(%) |
| --- | --- | --- | --- |
| **Age** |  |  |  |
| 18-39 | 369,190 (33.1) | 496,976 (35.0) | 548,230 (36.3) |
| 40 to 64 | 532,453 (42.1) | 591,894 (41.7) | 617,225 (40.9) |
| 65 to 79 | 244,094 (19.3) | 254,422 (17.9) | 266,812 (17.7) |
| >80 | 70,825 (5.6) | 74,993 (5.3) | 76,995 (5.1) |
| **Biological Sex** |  |  |  |
| Female | 706,897 (55.9) | 800,705 (56.4) | 848,266 (56.2) |
| Male | 557,837 (44.1) | 617,850 (43.6) | 661,317 (43.8) |
| **Payer** |  |  |  |
| Commercial | 816,373 (64.5) | 785,998 (55.4) | 766,183 (50.8) |
| Medicaid | 183,163 (14.5) | 361,613 (25.5) | 475,104 (28.2) |
| Medicare Advantage | 245,272 (19.4) | 270,944 (19.1) | 268,296 (21.1) |
| **Region** |  |  |  |
| Central | 254,754 (17.8) | 245,912 (17.3) | 258,587 (17.1) |
| Eastern | 274,952 (21.7) | 356,974 (25.2) | 369,590 (24.5) |
| Northern | 338,054 (26.7) | 316,810 (22.3) | 368,476 (24.4) |
| Northwest | 186,569 (14.7) | 234,413 (16.4) | 243,612 (16.0) |
| Southwest | 220,479 (17.4) | 264,446 (18.6) | 256,102 (17.0) |
| **Rurality** |  |  |  |
| Urban | 1,073,759 (84.9) | 1,198,679 (84.5) | 1,278,617 (84.7) |
| Rural | 190,975 (15.1) | 219,876 (15.5) | 230,966 (15.3) |

Of the NSAID prescription claims filled by patients with CKD, HF, and/or HTN during 2019-2021, 80.1% were categorized as high-risk pre-pandemic, with 83.3% high-risk during-pandemic. High-risk NSAIDs prescribing rates were greater in females vs. males (60.4 vs. 48.3 prescriptions/1000 patients) and in patients ages 40-64 and 65-79 years (75.6 and 72.8 prescriptions/1000 patients) vs. those 80+ and 18-39 years (55.8 and 12.0 prescriptions/1000 patients) (**Supplemental Files 1 and 2**). Extrapolated to the statewide population of insured adults, 5,841,000 high-risk NSAID prescriptions were filled by Virginians during 2019-2021.

### High-Risk NSAID Prescribing in Rural vs. Non-Rural Virginia

Throughout 2019-2021, unadjusted high-risk NSAID prescribing rates were 87.3% higher in rural vs. non-rural areas (92.8 vs. 49.6 prescriptions/1000 patients). Our model indicated a significant effect of rurality (IRR: 1.594 [95% CI: 1.408, 1.803], p<.001), as well as a significant interaction (p=.007) between rurality and biological sex (**FIGURE 1**). This interaction resulted in 28.5% greater predicted high-risk prescribing in females relative to males in non-rural areas, but only 12.1% higher in rural areas. The interaction between rurality and age was non-significant (**TABLE 2, Supplemental File 3**).

**FIGURE 1.**
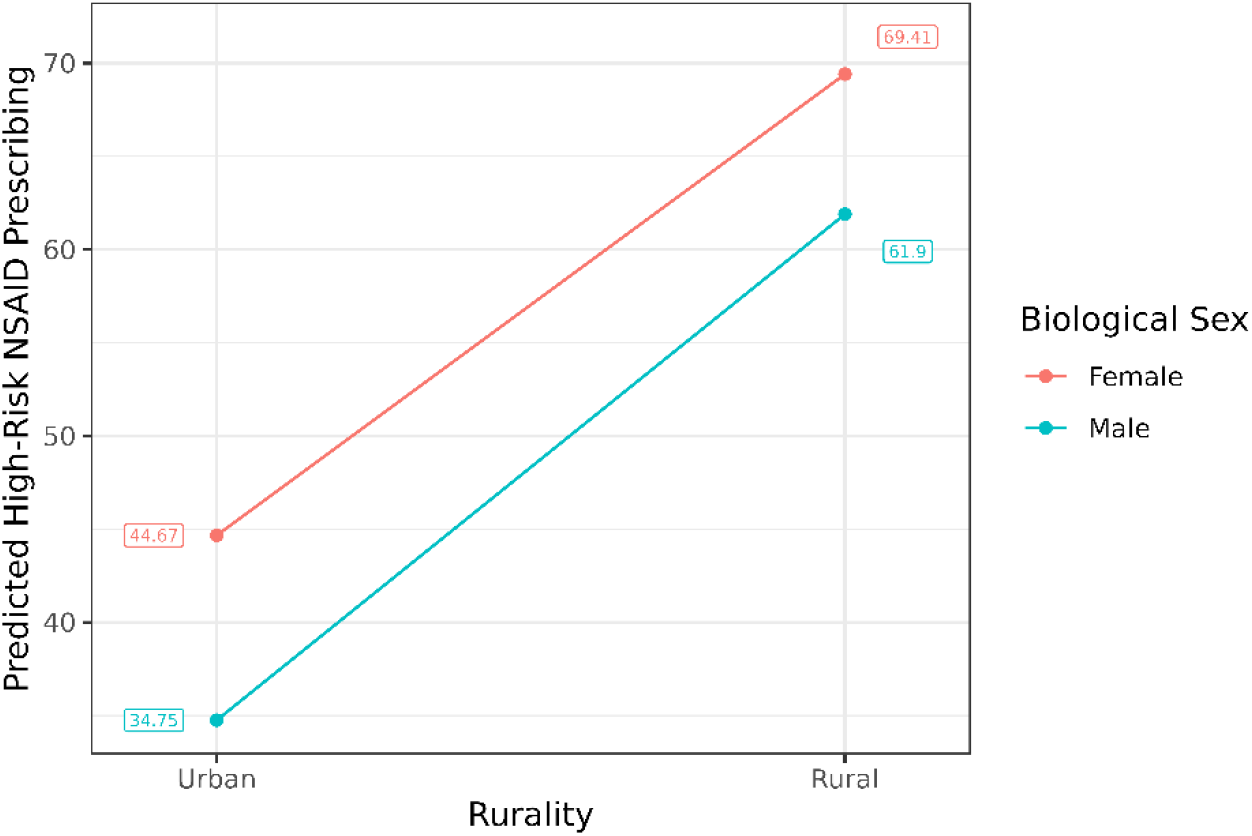
Interaction Plot: Rurality X Biological Sex for High-Risk NSAID Prescribing in Patients with Chronic Kidney Disease, Heart Failure, and/or Hypertension in Virginia in 2019-2021. This figure shows the significant (p=.007) moderating role of biological sex on the relationship between rurality and high-risk NSAID prescribing. Predicted marginal means are shown, calculated using group proportions within the study sample.

**Table 2.**
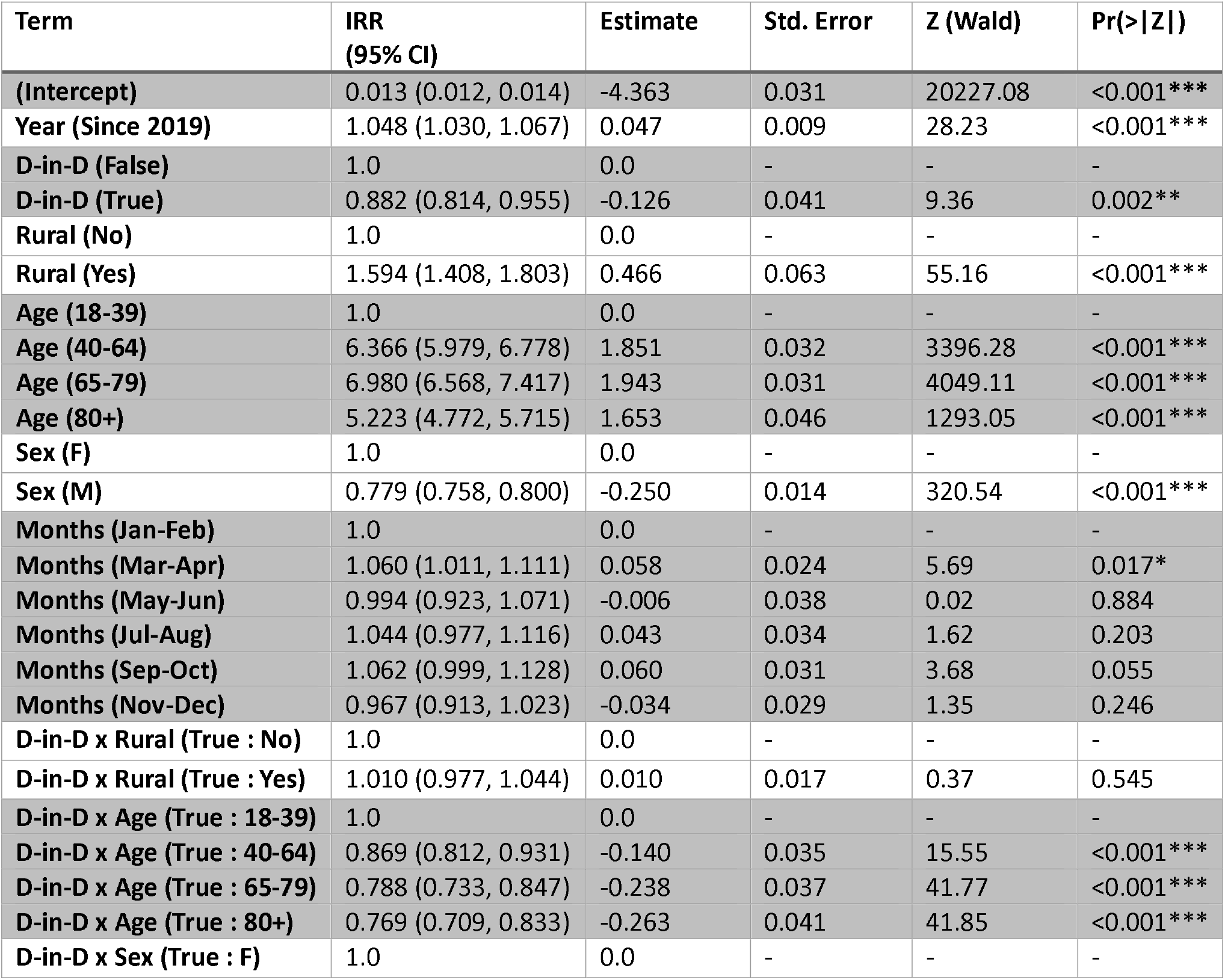
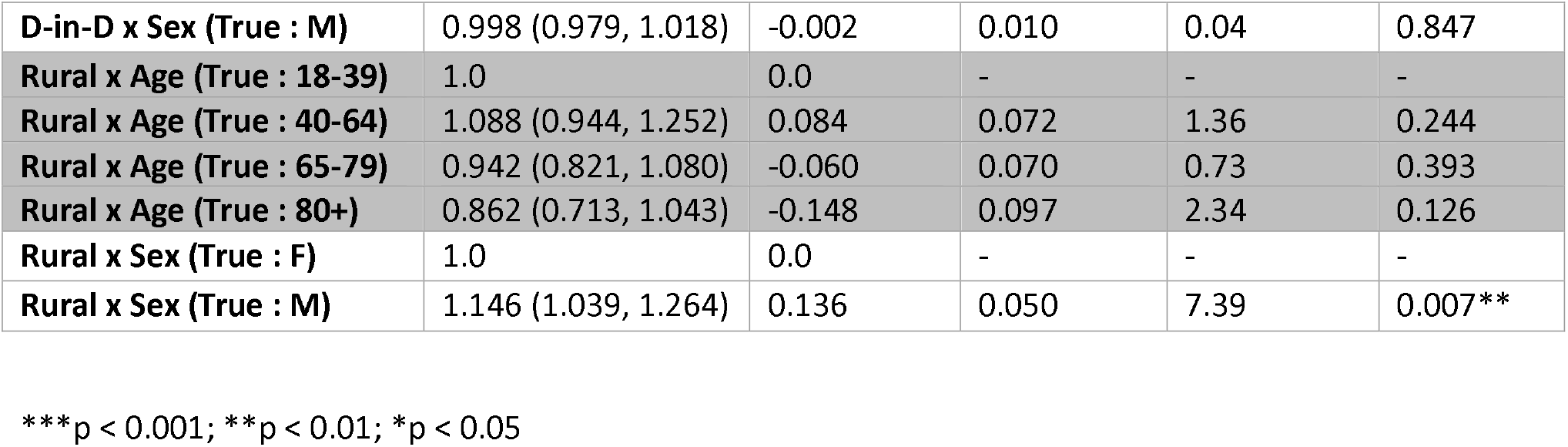
Poisson Regression Effects for Modeling Predicted High-Risk NSAID Prescribing among Patients with Chronic Kidney Disease, Heart Failure, and/or Hypertension in Virginia during March 1, 2020 to December 31, 2021.

| Term | IRR (95% CI) | Estimate | Std. Error | Z (Wald) | Pr(>\|Z\|) |
| --- | --- | --- | --- | --- | --- |
| (Intercept) | 0.013 (0.012, 0.014) | -4.363 | 0.031 | 20227.08 | <0.001*** |
| Year (Since 2019) | 1.048 (1.030, 1.067) | 0.047 | 0.009 | 28.23 | <0.001*** |
| D-in-D (False) | 1.0 | 0.0 | - | - | - |
| D-in-D (True) | 0.882 (0.814, 0.955) | -0.126 | 0.041 | 9.36 | 0.002** |
| Rural (No) | 1.0 | 0.0 | - | - | - |
| Rural (Yes) | 1.594 (1.408, 1.803) | 0.466 | 0.063 | 55.16 | <0.001*** |
| Age (18-39) | 1.0 | 0.0 | - | - | - |
| Age (40-64) | 6.366 (5.979, 6.778) | 1.851 | 0.032 | 3396.28 | <0.001*** |
| Age (65-79) | 6.980 (6.568, 7.417) | 1.943 | 0.031 | 4049.11 | <0.001*** |
| Age (80+) | 5.223 (4.772, 5.715) | 1.653 | 0.046 | 1293.05 | <0.001*** |
| Sex (F) | 1.0 | 0.0 | - | - | - |
| Sex (M) | 0.779 (0.758, 0.800) | -0.250 | 0.014 | 320.54 | <0.001*** |
| Months (Jan-Feb) | 1.0 | 0.0 | - | - | - |
| Months (Mar-Apr) | 1.060 (1.011, 1.111) | 0.058 | 0.024 | 5.69 | 0.017* |
| Months (May-Jun) | 0.994 (0.923, 1.071) | -0.006 | 0.038 | 0.02 | 0.884 |
| Months (Jul-Aug) | 1.044 (0.977, 1.116) | 0.043 | 0.034 | 1.62 | 0.203 |
| Months (Sep-Oct) | 1.062 (0.999, 1.128) | 0.060 | 0.031 | 3.68 | 0.055 |
| Months (Nov-Dec) | 0.967 (0.913, 1.023) | -0.034 | 0.029 | 1.35 | 0.246 |
| D-in-D x Rural (True : No) | 1.0 | 0.0 | - | - | - |
| D-in-D x Rural (True : Yes) | 1.010 (0.977, 1.044) | 0.010 | 0.017 | 0.37 | 0.545 |
| D-in-D x Age (True : 18-39) | 1.0 | 0.0 | - | - | - |
| D-in-D x Age (True : 40-64) | 0.869 (0.812, 0.931) | -0.140 | 0.035 | 15.55 | <0.001*** |
| D-in-D x Age (True : 65-79) | 0.788 (0.733, 0.847) | -0.238 | 0.037 | 41.77 | <0.001*** |
| D-in-D x Age (True : 80+) | 0.769 (0.709, 0.833) | -0.263 | 0.041 | 41.85 | <0.001*** |
| D-in-D x Sex (True : F) | 1.0 | 0.0 | - | - | - |
| **D-in-D x Sex (True : M)** | 0.998 (0.979, 1.018) | -0.002 | 0.010 | 0.04 | 0.847 |
| **Rural x Age (True : 18-39)** | 1.0 | 0.0 | - | - | - |
| **Rural x Age (True : 40-64)** | 1.088 (0.944, 1.252) | 0.084 | 0.072 | 1.36 | 0.244 |
| **Rural x Age (True : 65-79)** | 0.942 (0.821, 1.080) | -0.060 | 0.070 | 0.73 | 0.393 |
| **Rural x Age (True : 80+)** | 0.862 (0.713, 1.043) | -0.148 | 0.097 | 2.34 | 0.126 |
| **Rural x Sex (True : F)** | 1.0 | 0.0 | - | - | - |
| **Rural x Sex (True : M)** | 1.146 (1.039, 1.264) | 0.136 | 0.050 | 7.39 | 0.007** |

\*\*\*p < 0.001; \*\*p < 0.01; \*p < 0.05

### The Impact of the COVID-19 Pandemic on High-Risk NSAID Prescribing in Virginia

Compared with pre-pandemic prescribing rates, unadjusted high-risk NSAID prescribing rates declined by 23.3% during 2020-2021 (64.8 to 49.7 prescriptions/1000 patients) (**FIGURE 2**). Our model indicated a significant pandemic (D-in-D) effect (IRR: 0.882 [95% CI: 0.814, 0.955], p=.002), as well as a significant interaction (p< .001) between the pandemic (D-in-D) and age such that the pandemic had a greater impact on high-risk NSAID prescribing in patients 40+ years compared with those ages 18-39 years 29.9%, and 31.6% for Virginians ages 18-39, 40-64, 65-79, and ≥80 years. The pandemic (D-in-D) and rurality as well as the pandemic (D-in-D) and biological sex interactions were non-significant (**TABLE 2, Supplemental File 4**). Model results for clinically-indicated NSAIDs are shown in **Supplemental Files 5-8**.

**FIGURE 2.**
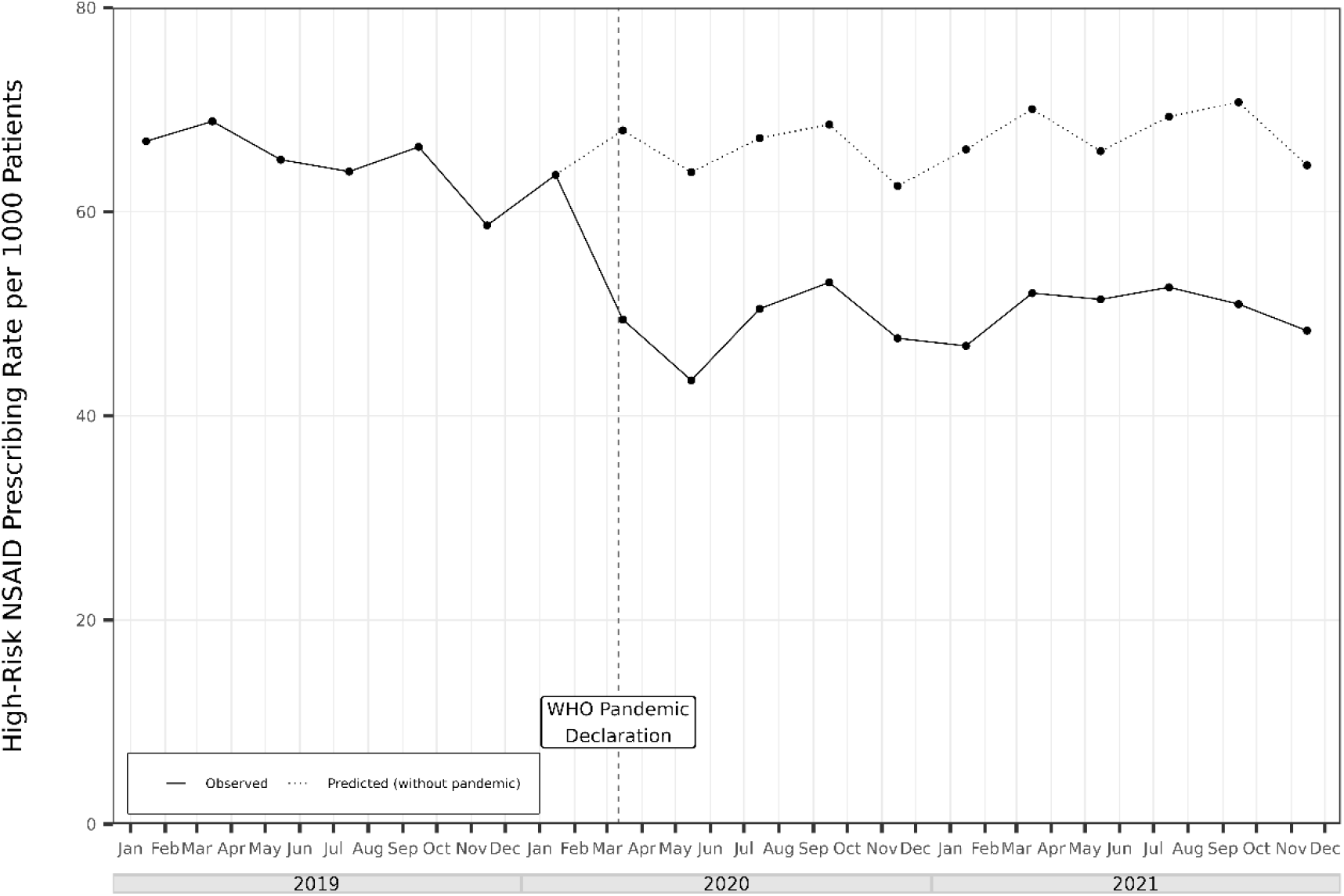
Observed vs. Predicted High-Risk Nonsteroidal Anti-Inflammatory Drug (NSAID) Prescribing Rates in Virginia in 2019-2021. Bi-monthly high-risk NSAID prescribing decreased from a mean of 64.8 prescriptions/1000 patients pre-pandemic to a mean of 49.7 prescriptions/1000 patients during-pandemic (a relative 23% decrease). NSAID= Nonsteroidal Anti-Inflammatory Drug WHO= World Health Organization. WHO declared the COVID-19 Pandemic on March 11, 2020. Pre-Pandemic= January 1, 2019 – February 28, 2020 During-Pandemic= March 1, 2020 – December 31, 2021

## DISCUSSION

Our analysis of claims data for >1 million Virginians identified a significant modest decrease in high-risk NSAID prescribing rates during the first two years of the COVID-19 pandemic. While this reduction in potentially harmful prescribing, which increased with age, is encouraging, the estimated overall volume of high-risk NSAIDs prescribed (∼5.8 million prescriptions statewide in 2019-2021) and the persistent disproportionate prescribing in rural areas (87.3% higher rates than non-rural) suggest opportunities to improve the quality of care for Virginians with chronic disease.

Our findings build upon the work of Levine et al.(15) who identified a small decline in high-risk NSAID prescribing during the first pandemic surge in 2020. Further research is needed to identify the mechanism for the observed decrease in high-risk NSAID prescribing rates. One possibility is reduced healthcare access during the pandemic. However, other studies have identified minimal impact of the pandemic on prescription medication utilization.(16,17) Clinicians may have improved adherence to evidence-based recommendations to limit or avoid NSAIDs in high-risk patients amidst the enhanced risk presented by the pandemic.

Public perception of the influence of NSAIDs on COVID-19 risk or prognosis may have led to decreased high-risk NSAID prescribing. Early pandemic reports suggested that NSAIDs suppressed immune function through enhancement of angiotensin-converting enzyme receptors (the entry point for SARS-CoV-2).(18,19) Although WHO and others reported no strong evidence linking NSAIDs to COVID-19,(20,21) the increased attention (e.g., 100% rise in related google searches in March 2020(22)) may have influenced prescribing behaviors.

Our study is among the first to identify disproportionately elevated rates of high-risk NSAID prescribing in rural vs. non-rural areas. The increased prevalence of CKD, HF, and HTN in rural areas offers only a partial potential explanation.(23,24) As rural Virginia has experienced some of the greatest impacts of the opioid epidemic, increased high-risk NSAID prescribing may reflect an unintended consequence of efforts to deprescribe opioids. Limited access to non-pharmacological pain therapies may also drive high-risk NSAID prescribing in rural areas.(10,11) It is unclear why biological sex differences (greater high-risk prescribing in females) were greater in non-rural vs. rural areas, but gender-based variation in NSAID use during the pandemic has also been reported elsewhere.(25)

### Limitations

Although our cohort included patients covered by three major insurers, findings may not generalize to patients covered by other insurers. Second, although our statistical model accounted for shifts in demographics during the study period, some changes in high-risk NSAID prescribing may have been misattributed to the pandemic. Third, claims data lack details about clinical interactions and decisionmaking that may influence prescribing rates. Finally, our data did not include details about prescription dose and quantity dispensed, which limited detection of prescribing variation.

### Conclusions

Among the most common medications used worldwide, NSAIDs pose cardiovascular and renal risk for patients with CKD, HF, and/or HTN. Although high-risk NSAID prescribing decreased significantly during the COVID-19 pandemic in Virginia, the high overall utilization (5.8 million prescriptions statewide in 2019-2021) and disproportionate utilization in rural areas suggest opportunity to enhance the quality of care in Virginia. Efforts to improve access to safe and effective pain management in rural areas may be particularly salient.

## Data Availability

All data produced are available online at the Virginia All Payer Claim Database

https://www.vhi.org/apcd/

## Acknowledgements

The authors thank Kyle Russell and Jillian Rider from Virginia Health Information for their expertise with the Virginia All-Payer Claims Database and for their generous collaboration, Beth Bortz from the Virginia Center for Health Innovation for her support of efforts to reduce low-value care in Virginia, and Emily Cox from the Carilion Clinic for her assistance with data management and manuscript preparation.

## Author Contribution Statement

Michelle S. Rockwell: conceptualization, methodology, data curation, writing-original draft, supervision, project administration, funding acquisition; Christopher Grubb: methodology, software, formal analysis, writing-review and editing, visualization; Jamie K. Turner: methodology, formal analysis, visualization; Matthew Vinson: investigation, writing-review and editing; Isaiah Yim: investigation, writing-review and editing; Alexandra L. Hanlon: methodology, formal analysis, supervision; John W. Epling: conceptualization, writing-review and editing, supervision

