## Supplemental Document for "Disproportionate High-Risk Nonsteroidal Anti-inflammatory Drug (NSAID) Prescribing in Rural Virginia"

**SUPPLEMENTAL FILE**

**TABLE OF CONTENTS**

**SUPPLEMENTAL FILE 1. Unadjusted Rates of High-Risk and Clinically-Indicated NSAID Prescribing among Patients with Chronic Kidney Disease, Heart Failure, and/or Hypertension in Virginia (2019-2021)**

**SUPPLEMENTAL FILE 2. Unadjusted High-Risk NSAID Prescribing Rates Pre- and During-Pandemic in Virginia Stratified by Rurality, Age, and Biological Sex**

**SUPPLEMENTAL FILE 3. Interaction Plot: Rurality X Age for High-Risk NSAID Prescribing in Patients with Chronic Kidney Disease, Heart Failure, and/or Hypertension in Virginia in 2019-2021**

**SUPPLEMENTAL FILE 4A and B. Interaction Plots: (A) Difference-in-Differences Effect (COVID-19 Pandemic) X Biological Sex and (B) Difference-in-Differences Effect X Rurality for High-Risk NSAID Prescribing in Patients with Chronic Kidney Disease, Heart Failure, and/or Hypertension in Virginia in 2019-2021**

**SUPPLEMENTAL FILE 5. Poisson Regression Effects for Predicting Clinically-Indicated NSAID Prescribing in Patients with Chronic Kidney Disease, Heart Failure, and/or Hypertension in Virginia (2019-2021)**

**SUPPLEMENTAL FILE 6A and B. Interaction Plots: (A) Rurality X Age and (B) Rurality X Sex for Clinically-Indicated NSAID Prescribing in Patients with Chronic Kidney Disease, Heart Failure, and/or Hypertension in Virginia in 2019-2021**

**SUPPLEMENTAL FILE 7. Interaction Plot: Difference-in-Differences Effect (COVID-19 Pandemic) X Age for Clinically-Indicated NSAID Prescribing in Patients with Chronic Kidney Disease, Heart Failure, and/or Hypertension in Virginia in 2019-2021**

**SUPPLEMENTAL FILE 8A and B. Interaction Plots: (A) Difference-in-Differences Effect (COVID-19 Pandemic) X Biological Sex and (B) Difference-in-Differences Effect (COVID-19 Pandemic) X Rurality for Clinically-Indicated NSAID Prescribing in Patients with Chronic Kidney Disease, Heart Failure, and/or Hypertension in Virginia in 2019-2021**

**SUPPLEMENTAL FILE 1. Unadjusted Rates of High-Risk and Clinically-Indicated NSAID Prescribing among Patients with Chronic Kidney Disease, Heart Failure, and/or Hypertension in Virginia (2019-2021)**

| **Year** | **Bi-monthly period** | **High-Risk**  **(prescription claims/1000 patients)** | **Clinically-Indicated**  **(prescription claims/1000 patients)** |
| --- | --- | --- | --- |
| **2019** | **Jan-Feb** | 66.9 | 14.8 |
|  | **Mar-Apr** | 68.9 | 15.2 |
|  | **May-Jun** | 65.1 | 15.4 |
|  | **Jul-Aug** | 64.0 | 16.1 |
|  | **Sept-Oct** | 66.4 | 17.5 |
|  | **Nov-Dec** | 58.7 | 17.1 |
|  |  |  | 24.0 |
| **2020** | **Jan-Feb** | 63.6 | 16.8 |
|  | **Mar-Apr** | 49.5 | 11.9 |
|  | **May-Jun** | 43.5 | 10.6 |
|  | **Jul-Aug** | 50.5 | 11.5 |
|  | **Sept-Oct** | 53.1 | 12.9 |
|  | **Nov-Dec** | 47.6 | 11.8 |
|  |  |  | 9.8 |
| **2021** | **Jan-Feb** | 46.9 | 9.2 |
|  | **Mar-Apr** | 52.0 | 8.7 |
|  | **May-Jun** | 51.4 | 8.2 |
|  | **Jul-Aug** | 52.6 | 8.2 |
|  | **Sept-Oct** | 51.0 | 8.6 |
|  | **Nov-Dec** | 48.4 | 8.7 |

**SUPPLEMENTAL FILE 2. Unadjusted High-Risk NSAID Prescribing Rates Pre- and During-Pandemic in Virginia Stratified by Rurality, Age, and Biological Sex**

|  | **Pre-Pandemic**  (prescriptions/1000 patients) | **During-Pandemic**  (prescriptions/1000 patients) | **% change from Pre-Pandemic to During-Pandemic** |
| --- | --- | --- | --- |
| **Overall** | 64.8 | 49.7 | -23.3% |
| **Rural** | 110.4 | 83.0 | -24.8% |
| **Urban** | 58.1 | 45.0 | -22.6% |
| **18 to 39 years** | 11.5 | 12.2 | +5.5% |
| **40 to 64 years** | 81.3 | 68.0 | -16.3% |
| **65 to 79 years** | 90.3 | 66.8 | -26.0% |
| **80+ years** | 68.0 | 48.5 | -28.8% |
| **Female** | 71.6 | 54.4 | -24.0% |
| **Male** | 56.5 | 43.8 | -22.4% |
| **18 to 39 years- Rural** | 22.0 | 17.9 | -18.5% |
| **18 to 39 years- Urban** | 10.6 | 11.6 | +9.7% |
| **40 to 64 years- Rural** | 141.2 | 111.2 | -21.3% |
| **40 to 64 years- Urban** | 73.0 | 62.0 | -15.1% |
| **65 to 79 years- Rural** | 128.1 | 99.7 | -22.2% |
| **65 to 79 years- Urban** | 82.8 | 60.8 | -26.6% |
| **80+ years- Rural** | 88.6 | 67.1 | -24.4% |
| **80+ years- Urban** | 63.8 | 44.9 | -29.7% |
| **Female- Rural** | 116.2 | 87.4 | -24.8% |
| **Male- Rural** | 103.6 | 77.6 | -25.0% |
| **Female- Urban** | 65.1 | 49.8 | -23.5% |
| **Male- Urban** | 49.5 | 38.9 | -21.3% |
| **18 to 39 years- Female** | 12.8 | 14.1 | +10.8% |
| **18 to 39 years- Male** | 10.2 | 9.8 | -4.0% |
| **40 to 64 years- Female** | 87.8 | 73.5 | -16.3% |
| **40 to 64 years- Male** | 74.1 | 61.7 | -16.7% |
| **65 to 79 years- Female** | 99.9 | 73.5 | -26.4% |
| **65 to 79 years- Male** | 77.7 | 58.1 | -25.2% |
| **80+ years- Female** | 76.1 | 54.2 | -28.8% |
| **80+ years- Male** | 53.7 | 38.5 | -28.2% |
| **18 to 39 years- Female-Rural** | 21.2 | 18.6 | -12.6% |
| **18 to 39 years- Male-Rural** | 22.8 | 17.2 | -24.6% |
| **40 to 64 years- Female-Rural** | 141.0 | 112.6 | -20.1% |
| **40 to 64 years- Male-Rural** | 141.4 | 109.6 | -22.5% |
| **65 to 79 years- Female-Rural** | 140.2 | 109.4 | -22.0% |
| **65 to 79 years- Male-Rural** | 112.9 | 87.7 | -22.3% |
| **80+ years- Female-Rural** | 98.8 | 75.2 | -23.9% |
| **80+ years- Male-Rural** | 70.4 | 52.9 | -25.0% |
| **18 to 39 years- Female-Urban** | 12.0 | 13.7 | +14.2% |
| **18 to 39 years- Male-Urban** | 9.0 | 9.0 | +0.3% |
| **40 to 64 years- Female-Urban** | 80.5 | 68.1 | -15.4% |
| **40 to 64 years- Male-Urban** | 64.5 | 54.8 | -15.0% |
| **65 to 79 years- Female-Urban** | 92.1 | 67.1 | -27.1% |
| **65 to 79 years- Male-Urban** | 70.6 | 52.5 | -25.6% |
| **80+ years- Female-Urban** | 71.4 | 50.1 | -29.8% |
| **80+ years- Male-Urban** | 50.3 | 35.8 | -28.8% |

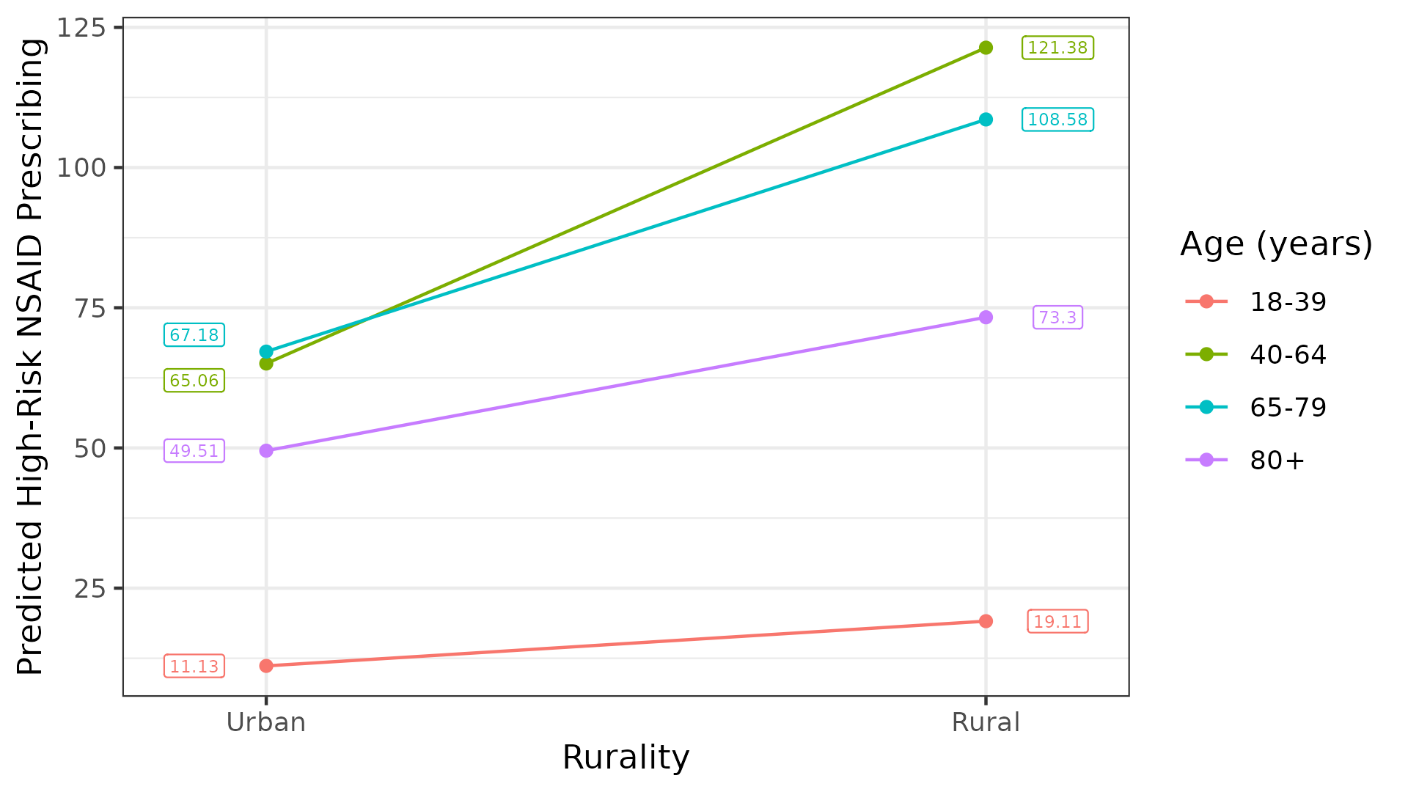

**SUPPLEMENTAL FILE 3. Interaction Plot: Rurality X Age for High-Risk NSAID Prescribing in Patients with Chronic Kidney Disease, Heart Failure, and/or Hypertension in Virginia in 2019-2021**

Caption: This figure shows the nonsignificant (p=.244, .393, and .126) moderating role of age group on the relationship between rurality and high-risk NSAID prescribing. Expected marginal means are shown, calculated using group proportions within the study sample.

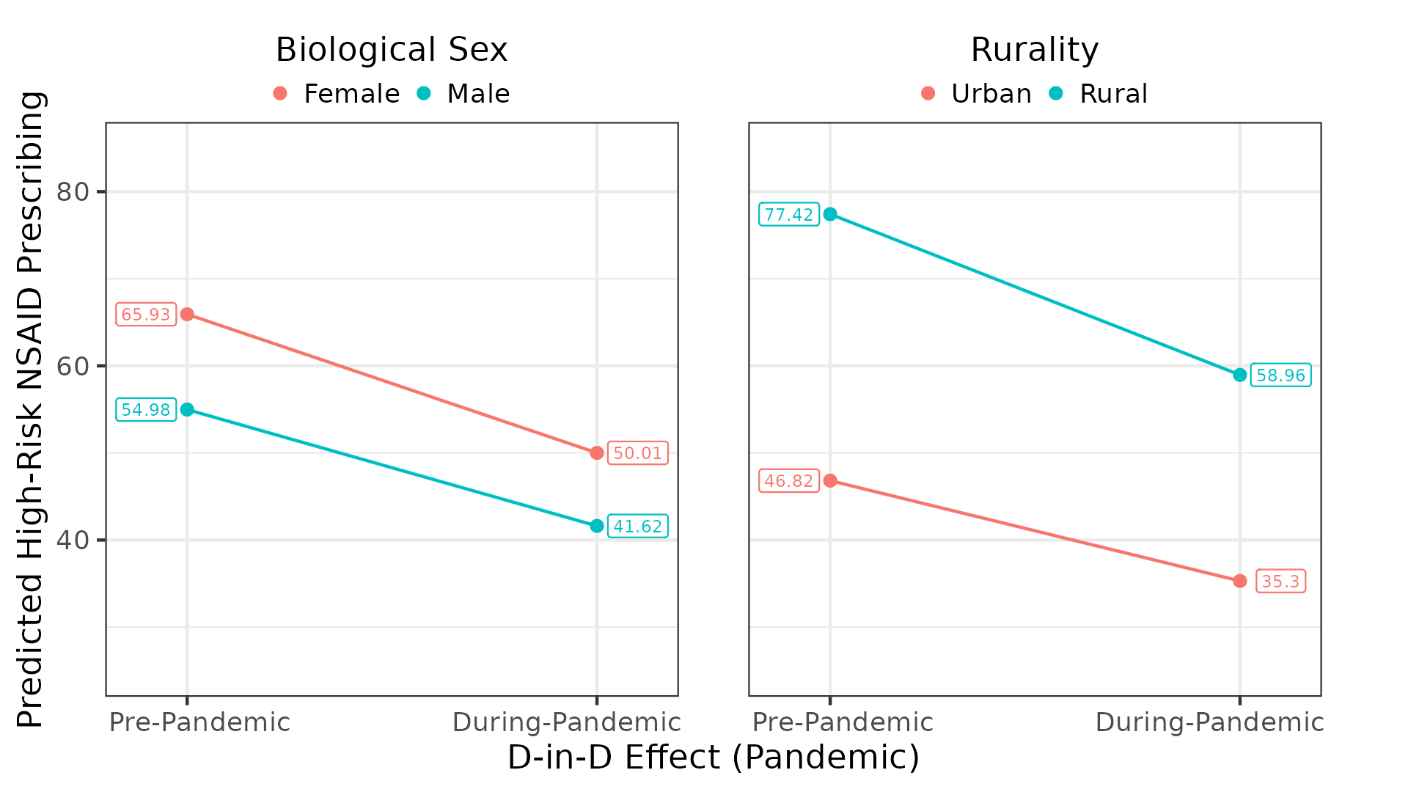

**SUPPLEMENTAL FILE 4A and 4B. Interaction Plots: (A) Difference-in-Differences Effect (COVID-19 Pandemic) X Biological Sex and (B) Difference-in-Differences Effect X Rurality for High-Risk NSAID Prescribing in Patients with Chronic Kidney Disease, Heart Failure, and/or Hypertension in Virginia in 2019-2021**

Caption: Figure A shows the nonsignificant (p=.847) moderating role of biological sex on the relationship between the pandemic and high-risk NSAID prescribing. Figure B shows the nonsignificant (p=.545) moderating role of rurality on the relationship between the pandemic and high-risk NSAID prescribing. Expected marginal means are shown, calculated using group proportions within the study sample.

**SUPPLEMENTAL FILE 5. Poisson Regression Effects for Predicting Clinically-Indicated NSAID Prescribing in Patients with Chronic Kidney Disease, Heart Failure, and/or Hypertension in Virginia (2019-2021)**

| Term | IRR (95% Conf. Interval) | Estimate | Std. Error | Z (Wald) | Pr(>\|Z\|) |
| --- | --- | --- | --- | --- | --- |
| (Intercept) | 0.001 (0.001, 0.001) | -6.759 | 0.048 | 19991.73 | <0.001*** |
| Year (Since 2019) | 0.911 (0.846, 0.982) | -0.093 | 0.038 | 6.14 | 0.013* |
| D-in-D (False) | 1.0 | 0.0 | - | - | - |
| D-in-D (True) | 0.631 (0.459, 0.866) | -0.461 | 0.162 | 8.05 | 0.005** |
| Rural (No) | 1.0 | 0.0 | - | - | - |
| Rural (Yes) | 1.716 (1.153, 2.555) | 0.540 | 0.203 | 7.09 | 0.008** |
| Age (18-39) | 1.0 | 0.0 | - | - | - |
| Age (40-64) | 15.074 (13.193, 17.224) | 2.713 | 0.068 | 1609.49 | <0.001*** |
| Age (65-79) | 23.618 (21.539, 25.897) | 3.162 | 0.047 | 4551.70 | <0.001*** |
| Age (80+) | 37.003 (32.577, 42.031) | 3.611 | 0.065 | 3069.05 | <0.001*** |
| Sex (F) | 1.0 | 0.0 | - | - | - |
| Sex (M) | 0.765 (0.688, 0.850) | -0.268 | 0.054 | 24.98 | <0.001*** |
| Months (Jan-Feb) | 1.0 | 0.0 | - | - | - |
| Months (Mar-Apr) | 0.967 (0.899, 1.039) | -0.034 | 0.037 | 0.86 | 0.354 |
| Months (May-Jun) | 0.912 (0.852, 0.977) | -0.092 | 0.035 | 7.09 | 0.008** |
| Months (Jul-Aug) | 0.958 (0.882, 1.040) | -0.043 | 0.042 | 1.08 | 0.298 |
| Months (Sep-Oct) | 1.029 (0.952, 1.113) | 0.0290 | 0.040 | 0.56 | 0.456 |
| Months (Nov-Dec) | 0.971 0.902, 1.047) | -0.029 | 0.038 | 0.59 | 0.443 |
| D-in-D x Rural (True : No) | 1.0 | 0.0 | - | - | - |
| D-in-D x Rural (True : Yes) | 1.132 (1.061, 1.208) | 0.124 | 0.033 | 14.16 | <0.001*** |
| D-in-D x Age (True : 18-39) | 1.0 | 0.0 | - | - | - |
| D-in-D x Age (True : 40-64) | 1.174 (0.869, 1.584) | 0.160 | 0.153 | 1.10 | 0.295 |
| D-in-D x Age (True : 65-79) | 1.141 (0.847, 1.537) | 0.132 | 0.152 | 0.75 | 0.387 |
| D-in-D x Age (True : 80+) | 1.058 (0.785, 1.425) | 0.056 | 0.152 | 0.14 | 0.713 |
| D-in-D x Sex (True : F) | 1.0 | 0.0 | - | - | - |
| D-in-D x Sex (True : M) | 1.044 (1.000, 1.090) | 0.043 | 0.022 | 4.05 | 0.044* |
| Rural x Age (True : 18-39) | 1.0 | 0.0 | - | - | - |
| Rural x Age (True : 40-64) | 0.694 (0.459, 1.050) | -0.365 | 0.211 | 3.00 | 0.083 |
| Rural x Age (True : 65-79) | 0.621 (0.416, 0.928) | -0.476 | 0.205 | 5.38 | 0.020* |
| Rural x Age (True : 80+) | 0.587 (0.380, 0.908) | -0.532 | 0.222 | 5.75 | 0.016* |
| Rural x Sex (True : F) | 1.0 | 0.0 | - | - | - |
| Rural x Sex (True : M) | 1.023 (0.912, 1.149) | 0.023 | 0.059 | 0.16 | 0.692 |

***p<.05, **p<.01, ***p<.001**

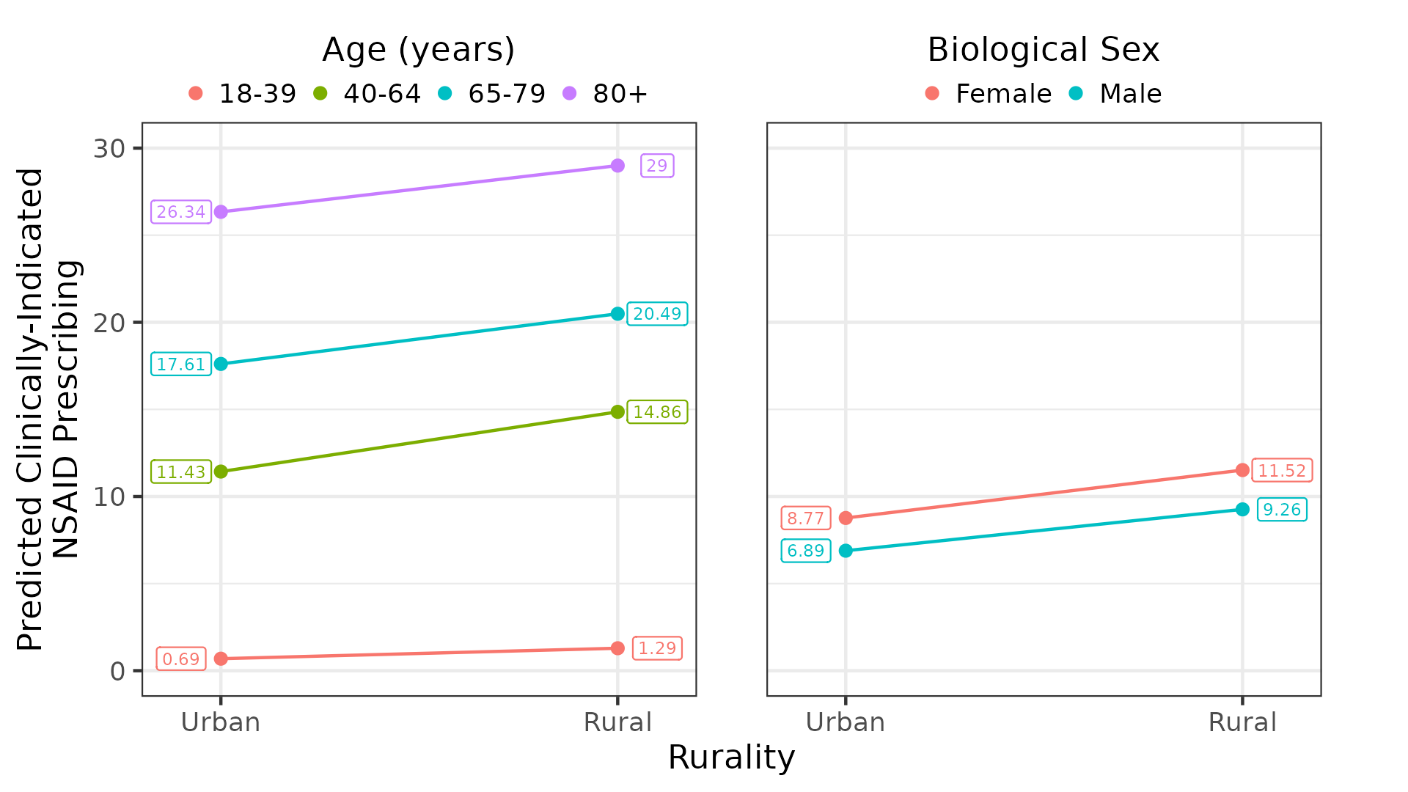

**SUPPLEMENTAL FILE 6A and B. Interaction Plots: (A) Rurality X Age and (B) Rurality X Sex for Clinically-Indicated NSAID Prescribing in Patients with Chronic Kidney Disease, Heart Failure, and/or Hypertension in Virginia in 2019-2021**

Caption: Figure A shows the significant (p=.083 for age 40-64, .020 for age 65-79, .016 for age 80+) moderating role of age on the relationship between rurality and clinically-indicated NSAID prescribing. Figure B shows the nonsignificant (p=.692) moderating role of biological sex on the relationship between rurality and clinically-indicated NSAID prescribing. Expected marginal means are shown, calculated using group proportions within the study sample.

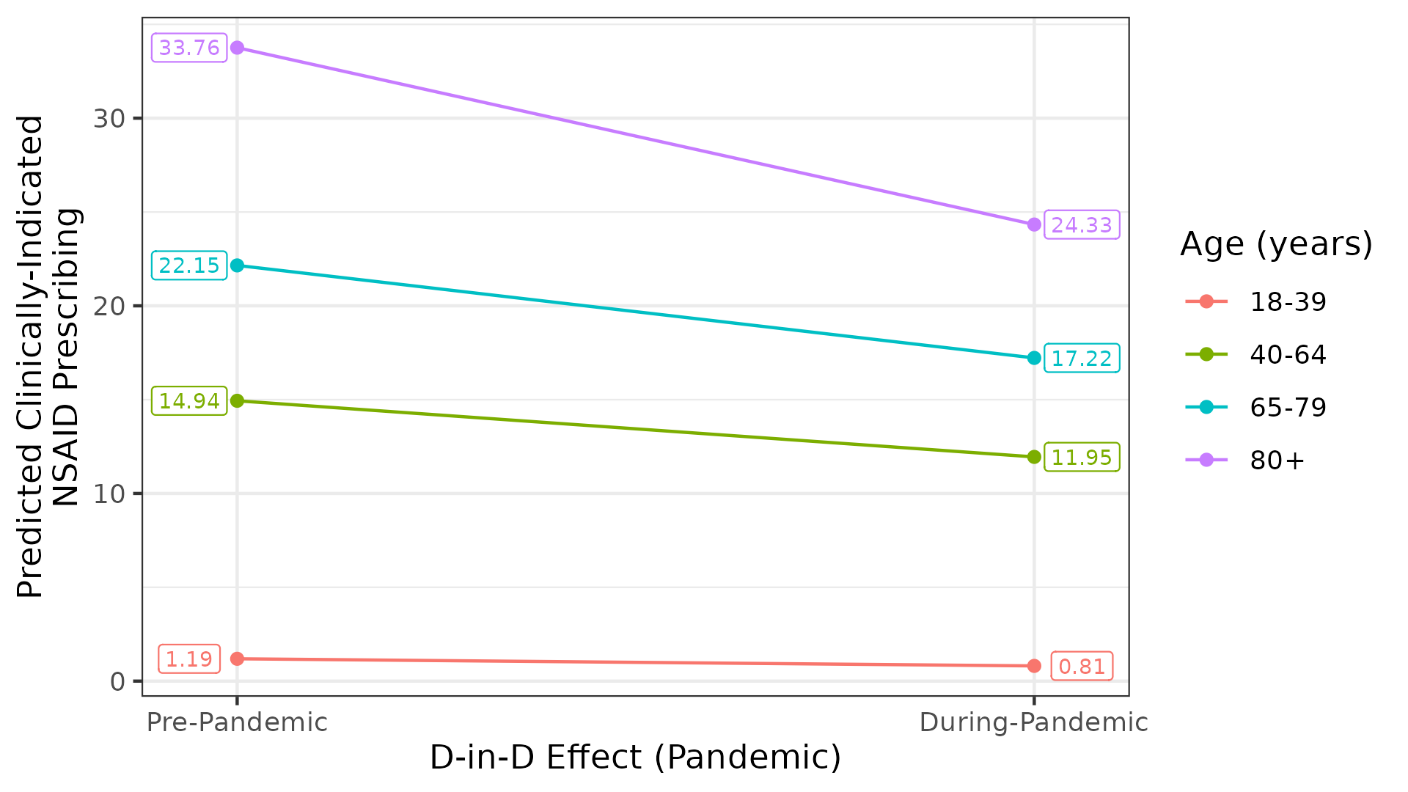

**SUPPLEMENTAL FILE 7. Interaction Plot: Difference-in-Differences Effect (COVID-19 Pandemic) X Age for Clinically-Indicated NSAID Prescribing in Patients with Chronic Kidney Disease, Heart Failure, and/or Hypertension in Virginia in 2019-2021**

Caption: This figure shows the nonsignificant (p=.295 for age 40-64, .387 for age 65-79, .713 for age 80+) moderating role of age group on the relationship between the pandemic and clinically-indicated NSAID utilization. Expected marginal means are shown, calculated using group proportions within the study sample.

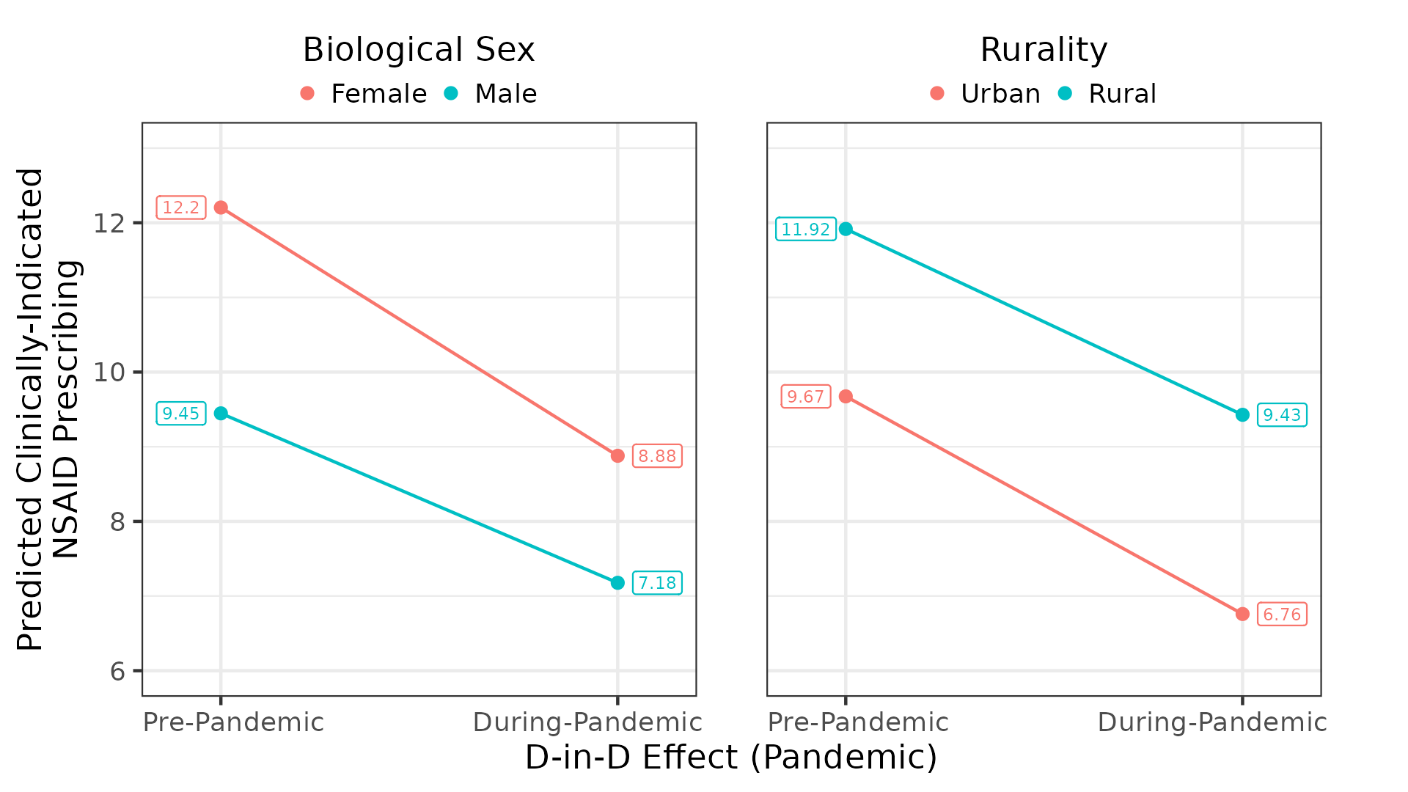

**SUPPLEMENTAL FILE 8A and B. Interaction Plots: (A) Difference-in-Differences Effect (COVID-19 Pandemic) X Biological Sex and (B) Difference-in-Differences Effect (COVID-19 Pandemic) X Rurality for Clinically-Indicated NSAID Prescribing in Patients with Chronic Kidney Disease, Heart Failure, and/or Hypertension in Virginia in 2019-2021**

Caption: Figure A shows the significant (p=.044) moderating role of biological sex on the relationship between the pandemic and clinically-indicated NSAID utilization. Figure B shows the significant (p<.001) moderating role of rurality on the relationship between the pandemic and clinically-indicated NSAID utilization. Expected marginal means are shown, calculated using group proportions within the study sample.

Our model indicated a significant interaction between the pandemic (D-in-D) and biological sex such that the pandemic had a significantly greater impact on clinically-indicated NSAID prescribing in female vs. male patients. Likewise, there was a significant interaction between the pandemic (D-in-D) and rurality such that the clinically-indicated NSAID prescribing rates in rural vs. urban Virginians were less affected. These interactions resulted in predicted during-pandemic marginal reductions of 27.2% in females vs 24.0% in males, as well as predicted marginal reductions of 20.9% for rural Virginians vs 30.1% for those living in urban Virginia. The pandemic (D-in-D) and age interaction was nonsignificant**.**
